# A systematic review of etiopathogenesis and treatment modalities for Moyamoya disease

**DOI:** 10.1101/2024.08.10.24311757

**Authors:** Ganesh Chilukuri, Amba Viswanathan, Amy Job, Vivek Joshi

## Abstract

**Introduction:** Moyamoya disease (MMD) is a rare and chronic cerebrovascular disease characterized by progressive stenosis or occlusion of the blood vessels within the terminal intracranial aspect of the internal carotid artery and the circle of Willis, leading to the compensatory development of a fragile collateral vessel network at the base of the brain. These vascular features are responsible for the recurrent ischemic and/or hemorrhagic strokes seen in affected patients. Numerous studies have attempted to clarify the clinical features of Moyamoya, including its etiopathology and interventions. In recent years, the development of neuroimaging and surgical techniques has enhanced the understanding of MMD in patients. The purpose of this review is to discuss the hypothesized etiopathogenesis, clinical manifestations, and the current possible treatment options available for Moyamoya disease.

**Methods:** The PRISMA protocol was utilized to perform an extensive literature search on Google Scholar, Scopus, Web of Science, and PubMed for articles about etiopathogenesis and clinical presentation of Moyamoya disease and the treatment protocols followed in different parts of the world. A comprehensive analysis was also conducted for original articles to better understand the possible clinical presentation and diagnostic criteria used.

**Results:** Based on the literature review, the RNF213 mutation, mitochondrial dysfunction, and misregulation of growth factors in endothelial cells are the most acceptable etiopathogenesis of MMD via their involvement in neurovascular inflammation.

**Conclusion:** The diagnostic test of choice is magnetic resonance angiography while direct & indirect revascularization surgeries are both effective and established treatments for managing symptoms of Moyamoya disease.

## 1. Introduction

Moyamoya disease (MMD) is a rare and chronic cerebrovascular disease characterized by progressive bilateral stenosis of the blood vessels within the terminal intracranial aspect of the internal carotid artery and the circle of Willis **^1–3^**. MMD leads to the compensatory development of a fragile collateral vessel network at the base of the brain. Moyamoya was first characterized in the 1960s when the compensatory vascular network was likened to a ‘puff of smoke’ in Japanese during cerebral angiography **^4^**. The incidence rate of MMD is high in Eastern and Southern Asia with the greatest percentage of reported cases originating from China, Japan, and Korea **^5^**. While the incidence of Moyamoya is low in European and North American populations **^6, 8^**, the number of patients diagnosed with MMD in these regions is seeing an upward trend **^7–8^**. The strong prevalence of Moyamoya disease in East Asian populations compared to other regions indicates support for a genetic trait associated with the disease. Much of the current literature demonstrates a difference in MMD prevalence based on biological sex with more females being affected than males **^5^**, but some Chinese literature cites an equal sex ratio regarding the prevalence of Moyamoya **^5, 9^**. Regardless, these findings suggest the role of sex differences in the clinical characteristics of Moyamoya, and Chinese cohorts may have differing presentations compared to other global regions. Furthermore, there is a global variation in the age of onset and diagnosis with general peaks of presentation in childhood and mid-to-late adulthood **^9–10^**. Due to this bimodal distribution, the average age of patients exhibiting Moyamoya clinical presentation is cited to be in the early-to-mid 30s **^3, 5, 9^**.

The vascular features of MMD result in a reduction of blood flow to the brain and lead to numerous potential clinical manifestations, including transient ischemic attacks (TIA), ischemic stroke, hemorrhagic stroke, epilepsy, infarction, headaches, cognitive impairment, and asymptomatic presentation **^11–12^**. Thus, MMD is described as a classic example of a hemodynamic cause of stroke. Clinically, children with MMD often present with ischemic attacks while adults exhibit either ischemic or hemorrhagic events **^2, 13–14^**. Considering the detrimental nature of these neurovascular symptoms, numerous studies have attempted to clarify the features of Moyamoya disease. Regardless, there is no confirmed etiopathology or cure for MMD, but in recent years, the development of neuroimaging and surgical techniques has enhanced our understanding of MMD in terms of potential pathophysiology and treatment modalities. As such, the objective of this review is to discuss the hypothesized etiopathogenesis, diagnostic methods, and available treatment options for Moyamoya disease.

## 2. Methods

This systematic literature review follows the guidelines determined by the Preferred Reporting Items for Systematic Reviews and Meta-Analyses (PRISMA) protocol. For this literature review, a comprehensive search was conducted in several databases including Worldwide Science, Scopus, Google Scholar, and PubMed. Prior to the literature screening process, the protocol was submitted and registered with the International Prospective Register of Systematic Reviews (PROSPERO ID: 488793).

### Selection Criteria

Studies included in this systematic review met these inclusion criteria,1) cross-sectional/observational studies examining the etiology and pathogenesis of Moyamoya disease, 2) Human studies conducted as prospective or Randomized Controlled Trial (RCT), 3) the research articles published between January 01, 2018, to October 01, 2023, and 4) full text articles, 5) published in the English language. 6) articles describing the diagnostic criteria and treatment modalities used in Moyamoya disease.

### Selection Strategy

A comprehensive literature search was conducted on October 11, 2023, using the electronic databases (PubMed, Worldwide Sciences, Google Scholar, and Scopus). Following search terms and keywords were utilized for the literature search:( (“Moyamoya Diseases” OR “Moyamoya Syndrome”) AND ((“Etiology” OR “Pathology” OR “Etiopathogenesis” OR “Mechanism”) AND (“TREATMENT” OR “THERAPEUTIC INTERVENTION” OR “MEDICATION” OR “PHARMACOLOGIC INTERVENTION” OR “DRUGS” OR “ALTERNATE THERAPIES” OR “SURGICAL INTERVENTIONS” OR “TREATMENT OPTIONS”)). Four authors (GC, AJ, AV and VJ) performed the literature search independently using the keywords for the articles to be included in this study. Those articles selected were screened for duplications or relevance, following which all the authors (GC, AV, AJ & VJ) reviewed the title and abstract independently for the research studies to be included. The article studies excluded did not meet the study objective or the inclusion criteria. Finally, the full texts of the selected articles were examined to determine the final exclusion for our systematic review by reaching a consensus with all the authors involved. We also excluded articles which were editorials, opinions, commentaries, and book chapters. Any study which was duplicated or involved the animal models was excluded (**Figure 1)**.

### Data Collection

A predefined table was utilized to extract the relevant information from each selected research study. The following information was extrapolated from each included research study: 1) Title, 2) First author, 3) Location/Country of study population, 4) Study design, 5) Etiopathogenesis, 6) Diagnostic criteria/findings, 7) Treatment modality used, 8) Results, 9) Conclusion.

### Data Analysis/Synthesis

Prior to this systematic literature review, an initial literature search was conducted in this selected study area which revealed a limited number of research studies and lack of comprehensive information about the Moyamoya disease. This limitation provided us the opportunity to conduct a qualitative analysis on the research hypothesis.

### Quality assessment

The quality of research studies and risk of any bias for the selected studies was performed by two independent reviewers (VJ and BMG) using the National Heart, Lung, and Blood Institute Quality Assessment Tool for Observational Cohort and Cross-Sectional Studies (Study Quality Assessment Tools; https://www.nhlbi.nih.gov/health-topics/study-qualityassessment-tools). Based on the predefined 14 item questionnaire, an overall rating was assigned to the studies as good, fair, and poor, which eliminated the risk of bias for the research studies included in the final systematic review.

## 3. Results

The database search identified 506 papers using the keywords. Initial screening for language, availability, and removal of duplicates eliminated 437 articles, leaving 69 studies for title/abstract screening. Three reviewers assessed the abstracts/titles of the papers to identify 25 articles total for the full-text review phase. 14 total articles met the eligibility criteria for inclusion (Figure 1). Additional information about each selected article is briefly summarized qualitatively; some articles overlapped in their discussions of treatments and etiopathogenesis **or** treatments and diagnostic measures (Tables 1-4).

Based on the most current literature, the most widely accepted hypothesized etiopathogenesis for MMD involves mechanisms surrounding mutations in the susceptibility gene (RNF213), endothelial mitochondrial dysfunction, and/or misregulation of growth factors in endothelial cells (ECs). Each of these mechanisms could be a plausible etiology resulting in MMD via their involvement in neurovascular inflammation. An alternative etiopathology that has been suggested is an upregulation of protein such as Filamin A in the dura mater leading to a variety of symptoms. Etiopathogenesis was also seen to vary when considering those with asymptomatic MMD. Particularly, a commonality of impaired expression of transfer RNA-derived small RNAs was found among this patient population. Risk factors for MMD were also found and included elevations in serum uric acid, triglycerides, and branched-chain amino acids (Figure 2).

Additionally, this study also found common clinical manifestations that patients with MMD can present with, including transient ischemic attacks (TIA), ischemic stroke, hemorrhagic stroke, epilepsy, infarction, headaches, cognitive impairment (including motor, sensory, and speech deficits), and/or no symptoms. When patients with similar symptoms and history present, the diagnostic test of choice has conventionally been a cerebral angiography using a catheter (Figure 3). However, more recently, magnetic resonance angiography has gained popularity. Additional imaging modalities including computerized tomography, positron emission tomography, EEG, and conventional magnetic resonance imaging are also used in the diagnosis of MMD (Figure 3).

Though no cure for Moyamoya disease currently exists there are a variety of treatment modalities including medication therapy aimed at symptomatic control and management (Figure 4). However, surgical treatment is the current most effective procedural management available for symptomatic control. The three main surgical options are direct revascularization, indirect revascularization, and combined revascularization (Figure 4). As each present with strengths and weaknesses, the current literature surrounding the establishment of a surgical modality that is most effective remains conflicting. However, overall direct & indirect revascularization surgeries are both effective and established treatments for managing symptoms of Moyamoya disease, and the literature recommends that surgical treatments be chosen based on the individual patient’s medical history and risk assessment.

## 4. Discussion

### 4.1. Etiopathogenesis

The causative mechanisms of Moyamoya disease are not confirmed; however, in this literature review, seven potential etiopathologies have been identified. The most plausible and accepted etiopathogenic mechanisms involve mutations in the susceptibility gene (RNF213), endothelial mitochondrial dysfunction, and misregulation of growth factors in endothelial cells (ECs) **^16, 18, 20–23, 28–29^**. An additional etiopathology is the upregulation of proteins in extracellular matrix organization–such as Filamin A–in the dura mater, leading to impaired ECM remodeling and hypermobility of vascular smooth muscle cells **^18, 30^**. Patients with asymptomatic MMD were revealed to have impaired expression of transfer RNA-derived small RNAs (tsRNAs), which normally regulate inflammation and neurovascular angiogenesis **^21^**. Additionally, elevated levels of serum uric acid (SUA), triglycerides, and branched-chain amino acids (BCAAs) are significant risk factors for MMD **^22, 29^**. Normal SUA is involved in lipid peroxidation pathways while BCAAs transduce the mTOR pathway, and impairments result in the generation of reductive oxygen species, inflammation, and angiogenic difficulties **^22, 29, 31–33^**. One hallmark of all these potential MMD pathologies is the presence of neuroinflammation **^23, 34^**.

#### 4.11. Susceptibility gene RNF213 and cerebral endothelial integrity

RNF213 was identified as a susceptibility gene for Moyamoya disease **^35–36^**, and recent developments have been made to elucidate the function of this gene in vascular development. Multiple single nucleotide polymorphisms (SNPs) have been found to increase the risk of MMD, indicating an autosomal dominant hereditary component to this cerebrovascular disease **^37^**. In particular, the p.R4810K variant predominated in East and South Asian populations while European/American patients exhibited other distinct variants to greater degrees of rarity **^16, 37–38^**. It is also important to note that specific mutations have been ascribed to certain clinical presentations such as asymptomology, ischemia, hemorrhage, or epilepsy and individual surgical outcomes **^39–42^**.

RNF213, also known as *Mysterin*, is a large protein consisting of 5,207 amino acids with a size of 591-kDa; it has both ATPase and E3 ligase activity. RNF213 is an important protein for lipid metabolism, inflammation, and ubiquitination ligase activity. Mutations associated with MMD are dominantly observed in the E3 ligase domain. The E3 enzyme engages in tagging unwanted or defective proteins via ubiquitination for digestion or disposal from cell **^43^**. SNPs related to Moyamoya RNF213 interfere with ubiquitin ligase activity, leading to either mechanistic inhibition or altered substrate binding, both of which result in accumulation of debris proteins **^44^**. This mechanism proposes an impairment of endothelial cell function due to decreased integrity of ECs, weakening of the blood-brain barrier (BBB), and elevated pro-inflammatory (leukocytic) response **^45–46^**. Specifically localized to vessels near the base of the brain, these genetic factors affecting BBB integrity and vascular angiogenesis point to the onset of MMD pathology. Additionally, mutated RNF213 may mediate pathogenesis via the Hippo pathway, resulting in overexpression of vascular endothelial growth factors and unnecessarily induced angiogenesis **^47^**.

#### 4.12. Endothelial mitochondrial dysfunction

Many studies highlight that patients with Moyamoya disease show lower levels of circulating endothelial colony-forming cells (ECFCs), previously known as endothelial progenitor cells (EPCs), which function to support and regenerate endothelial cells, especially in times of damage **^28–29, 48–49^**. However, some studies emphasize that increased levels of circulating ECFCs or EPCs are pathogenic markers of MMD, leading to hypermobility of ECs, mixed conditions of vascular occlusion, and abnormal angiogenesis **^50^**. Regardless of the patient population’s characteristics of ECFCs, it is accepted that the ECs, ECFCs, or EPCs are generally impaired, defective, or misregulated. One method for the altered functionality of ECs, immune cells, and/or vascular repair cells is mitochondrial dysfunction in these cells [28-29]. Several studies have elucidated that the EPFCs in patients with MMD revealed morphologically and functionally abnormal mitochondria **^28–29^**. Mitochondrial dysfunction and the subsequent disruptions in metabolic homeostasis can cause oxidative stress, calcium sequestration, CNS neuroinflammation, and cell impairment/death. These factors can weaken/occlude arteries by delaying vessel repair and forming compensatory micro-vessels, leading to risks of ischemic crisis, abnormal blood flow (resulting in seizures), and hemorrhage.

Many abnormal mechanisms (genetic and acquired) can lead to a disruption in mitochondrial activity, and research is still progressing to elucidate the specific mechanisms associated with MMD. First, BCAAs, which tend to exist at high levels in Moyamoya patients, transduce the mTOR pathway. The mTOR pathway–a regulator of cell growth and metabolism–also influences mitochondrial function, and disruptions result in excessive production of reactive oxygen species, leading to EC impairment [28, 55]. Another mechanism incorporates the lower expression of Coenzyme Q10B (CoQ10B, a mitochondrial electron carrier and antioxidant) seen in MMD patients; Jian, 2021 postulate the use of CoQ10 supplements for therapeutic management of Moyamoya symptoms **^29^**. Finally, mitochondrial function and many growth factors act in conjunction to facilitate proper development and action of endothelial, immune, and nervous system tissues.

#### 4.13. Misregulation of growth factors (GF) in endothelial cells

Numerous studies have demonstrated that patients with MMD have elevated levels of multiple cytokines–including but not limited to vascular endothelial growth factor (VEGF), basic fibroblast growth factor (bFGF), hepatocyte growth factor (HGF), cellular retinoic acid binding protein 1, and granulocyte-colony-stimulating factor–in the blood, cerebrospinal fluid, dura mater, and arachnoid mater **^20, 28^**. VEGF is a prominent angiogenic factor that has been linked to MMD presentation. The common pathology regarding this overexpression and misregulation of growth factors is described as a dysregulated proliferation and migration of ECs and smooth muscle cells, leading to intimal thickening that results in luminal stenosis in the internal carotid artery and the circle of Willis. The stenosis and occlusion point to a risk of ischemia due to decreased blood flow. Meanwhile, having too many circulating growth factors and inflammatory proteins can lead to the formation of unnecessary collateral vessels (Moyamoya vessels), which are aneurysmatic formations and risks for hemorrhage. It is worth mentioning that there is contention in the field that misregulation of growth factors may be a secondary association rather than causative of MMD. In other words, underlying genetic mutations or cellular dysfunction may be influencing the altered activity of various growth factors–either through an upregulated expression of GFs or downregulation of GF receptors **^41^**. Regardless, VEGF levels and GF receptor levels may play a valuable role in diagnostic and prognostic measures after surgical revascularization treatment.

### 4.2. Diagnostic Measures

As mentioned above, patients with MMD can present with transient ischemic attacks (TIA), ischemic stroke, hemorrhagic stroke, epilepsy, infarction, headaches, cognitive impairment (including motor, sensory, and speech deficits), and no symptoms **^14^**. After the presence and acute treatment of the early symptoms, physicians consult and collaborate to discover the underlying cause. Many neuroradiologic diagnostic tools have been developed to study cerebrovascular disorders like Moyamoya disease. The diagnostic test of choice is conventional cerebral angiography using a catheter; however, magnetic resonance angiography (MRA) has become increasingly popular due to being less invasive and still giving sensitive results **^1, 14, 24–25, 27^**. Computerized tomography, positron emission tomography, EEG, and conventional magnetic resonance imaging are additional neuroimaging tools that are useful for visualizing specific aspects of MMD presentation such as the locations of strokes/hemorrhage, damaged neurovascular structures, atypical brain activity, **^25, 27^**. To make a diagnosis of Moyamoya disease, imaging must show the following minimum findings: (i) visible stenosis/occlusion of terminal ICA or proximal ACA and/or MCA, (ii) presence of abnormal vascular networks near stenotic/occlusive lesions or in basal ganglia, (iii) bilaterality of findings i and ii. Finally, MMD vasculopathy is associated with other clinical disorders like neurofibromatosis type 1, Down syndrome, thyroid disease, cranial irradiation, sickle cell disease, and many autoimmune disorders; in these patients, the condition is termed Moyamoya Syndrome **^50^**.

### 4.3. Treatment Modalities

No known cure for Moyamoya disease exists, so the goals of treatment are management of symptoms, prevention of stroke/hemorrhage, lower risks of complications to surgery, and improvement of blood flow to the brain. First, medication therapy focuses on symptomatic control with antiplatelet therapy and anticoagulant drugs in conjunction with neurotropic medications being a popular choice **^17, 46^**. Additionally, if patients exhibit other symptoms such as epilepsy or cognitive impairment, corticosteroids, calcium-channel blockers, anti-seizure drugs, etc. are provided for symptom relief. While medication therapy provides short-term symptomatic control for patients with MMD (especially patients with mild presentation or those preparing for surgical treatment), its effectiveness remains unclear and controversial in the long run. Furthermore, endovascular treatment (EVT) has been shown to provide safe and effective embolization/stenting of intracranial aneurysms to improve blood flow; however, due to the location of MMD stenosis in the ICA and promotion of inflammation by EVT, the long-term clinical impacts remain controversial and need for further review **^48–49^**.

Surgical treatment remains the most effective symptom management procedure for patients with Moyamoya disease. Currently, there are three main surgical options for patients: direct revascularization, indirect revascularization, and combined revascularization. Direct revascularization commonly involves anastomosis of the superficial temporal artery to the middle cerebral artery (STA-MCA bypass) but also supports leptomeningeal to the anterior carotid artery (ACA) anastomosis **^27^**. Direct bypass surgery has been shown to reduce ischemic and hemorrhagic risk through the immediate enhancement of blood perfusion, but it remains a relatively difficult and invasive procedure. Meanwhile, indirect revascularization involves the overlay of blood-rich tissues directly over the brain’s surface using encephalo-duro-arterio- synangiosis (EDAS), encephalomyosynangiosis (EMS), burr holes, and omental transplants [D41-43]. The goal of indirect revascularization is the spontaneous, progressive ingrowth of new blood vessels from the overlayed scalp or muscle tissue. This procedure is easier to perform, non-invasive, and shows improvements in stroke though risk of hemorrhage persists. Finally, combined revascularization uses a combination of both direct and indirect revascularization procedures to provide instant and progressive improvements in blood flow to the brain. Usually, a failsafe after direct revascularization complications, combined revascularization is an effective treatment strategy, but due to the novelty of this procedure in treatment MMD patients, additional evidence is urgently needed. There is much controversy amongst scientists on the superiority of these three surgical procedures, but direct and indirect revascularization remains established and effective treatments with their own merits and demerits. Current research highlights conflicting results on which procedure is more useful for managing MMD. Significantly, indirect bypass may provide enhanced clinical benefits for pediatric MMD while direct bypass demonstrates significant improvements for adult patients **^17–20, 26–27, 40–44^**. Overall, the authors recommend utilizing direct and indirect modalities of revascularization depending on the individual patient’s medical history and risk assessment.

Patients with MMD are often recommended physical, occupational, educational, and/or psychological therapy to address the physical, emotional, and cognitive impacts of stroke/hemorrhage. Post-operative quality of life and daily functional outcomes can be significantly improved with psychological intervention and routine monitoring of symptoms, especially considering the chronic and progressive nature of Moyamoya disease **^26^**.

## 5. Conclusion

Based on the literature review conducted, there are several etiologies that could play a role in Moyamoya disease. The more widely accepted etiopathogeneses are mutations in the susceptibility gene (RNF213), endothelial mitochondrial dysfunction, and misregulation of growth factors in endothelial cells. The test of choice in diagnosing MMD is MRA. The most effective treatment modality for MMD has been shown to be surgical interventions; both direct revascularization surgery and indirect revascularization surgery are widely used to treat symptoms of MMD in patients. In the future, we would like to look further into the comparison between direct and indirect revascularization and conduct a retrospective chart review on patients who have undergone either surgical intervention to elucidate the advantages and disadvantages of both methods.

## Data Availability

All data produced in the present study are available upon reasonable request to the authors.

## Contribution

The authors GC, AV, AJ, and VJ conceptualized the study. GC, AV, AJ, and VJ wrote the first draft. GC, AJ, AV, and VJ analyzed and interpreted the data. All the authors (GC, VJ, AV, and AJ) contributed to its administration, discussion, conclusion, and critical revision. All authors approve the final manuscript for submission.

## Data availability

The data generated during the research and analysis are not available publicly but are available from the corresponding author on a reasonable request.

